# A Comparative Environmental Impact Analysis of Screening Tests for Colorectal Cancer

**DOI:** 10.1101/2025.01.14.25320553

**Authors:** Vivek A. Rudrapatna, Tzu An Wang, Parsia Vazirnia, Kaiyi Wang, Nathan Alhalel, Shadera Slatter, Gunnar Mattson, Amy Becker, Ching-Ying Oon, Shan Wang, William Karlon, Scott Pasternak, Cassandra L. Thiel, Seema Gandhi, Sean Woolen

## Abstract

**BACKGROUND:** Healthcare is a major contributor to global greenhouse gas emissions. Colorectal cancer (CRC) screening is one of the most widely used healthcare services in the US, indicated for approximately 134 million adults. Recommended screening options include fecal immunochemical tests (FITs) every year, CT colonographies (CTCs) every 5 years, or colonoscopies every 10 years. We compared the environmental impacts of these tests and identified opportunities for impact reduction.

**METHODS:** We conducted a comparative life cycle assessment of three CRC screening strategies at the University of California, San Francisco. We performed on site audits to document the materials and energy used for each screening test. We used the ReCiPe 2016 method to estimate the environmental impacts of these procedures, measured by global warming potential (GWP) and damage to human health. We estimated the 10-year cumulative impacts of each screening strategy using a Markov reward model. We accounted for model uncertainty using hierarchical Monte Carlo simulations.

**FINDINGS:** FIT-based screening had the lowest environmental impacts, with a roughly 20% margin of superiority over colonoscopies, and this finding was robust in sensitivity analyses. Across tests, the biggest cause of environmental harm was car-based transportation of patients and staff. Prioritizing FITs over screening colonoscopies in the US could enhance population health by roughly 5.2 million disability adjusted life years per decade. Transitioning to electric vehicles could reduce the GWP of all screening tests by 15-20%.

**INTERPRETATION:** Given the similar efficacy and safety of these tests, payors should prioritize FITs for low-risk patients. Government initiatives to decarbonize transportation, incentivize telehealth, and mandate environmental product declarations will help reduce the environmental impacts of healthcare more generally. Our results call for a closer look at resource-intensive preventative health strategies, which could result in more harm than good if applied to a low-risk population.

**FUNDING:** NIH, UCSF

## INTRODUCTION

According to the World Health Organization, climate change is “the single biggest health threat facing humanity”^1^. Ironically, healthcare services are a significant contributor to this crisis, responsible for nearly 10% of the US’s greenhouse gas emissions^2^. More concerningly, emissions due to US healthcare are rising faster than other sectors of the economy^2,3^. These trends call for an urgent look at high volume services in medicine, particularly those where comparable alternatives are available.

Colorectal cancer (CRC) screening is indicated for roughly 134 million adults in the US. The US Preventative Services Task Force (USPSTF) currently recommends multiple screening options for average-risk individuals. These include fecal immunochemical tests (FITs) every year, CT colonographies (CTCs) every 5 years, or colonoscopies every 10 years^4^. These tests have been extensively studied and are thought to have broadly similar efficacy (e.g. mortality benefit) and safety^5^.

As such, the predominance of any given screening test has largely been a function of economics. Screening colonoscopies have been popular in the US due to its history of fee-for-service reimbursement^6^. By contrast, FIT-based screening is common in regions with single-payor healthcare and organized public screening, including Europe, South America, and East Asia^7–9^.

While the cost-effectiveness of these screening strategies has been extensively studied, few studies have evaluated their environmental impact. There are no prior studies estimating the environmental impacts of FITs or CTCs. Moreover, there have not been any comparative studies incorporating life cycle assessments (LCAs), the gold-standard for estimating environmental impacts.

We performed a comparative LCA of three screening strategies recommended by the USPSTF: FITs, CTCs, and colonoscopies. We hypothesized that these otherwise comparable tests may differ in their impacts on climate change and future human health.

## METHODS

This study was conducted at the University of California, San Francisco (UCSF). It did not involve human subjects and thus was exempted from institutional review board oversight.

### STUDY DESIGN

We performed an LCA to compare the environmental impacts of CRC screening by three different modalities. Our functional unit of study was screening one average risk patient (age 45) over 10 years. The study team included physicians, hospital administrators, and a healthcare LCA expert.

To define study boundaries, we first classified component processes as either variable (i.e. opportunity) or fixed costs. Opportunity costs for colonoscopies and CTCs included bowel preparation, as well as transportation of patients to/from testing facilities. Fixed costs included the energy consumed by colonoscope ventilation cabinets (required to be continuously on irrespective of use, per the manufacturer). Staff transportation for FITs was also considered a fixed cost: under counterfactual scenarios we assumed staff would still travel to run other laboratory tests within the hospital.

Our study boundary included all opportunity costs: environmental harms that could be avoided (or incurred) following a change in screening preferences. We excluded all fixed costs. For example, we ignored the baseline energy for heating, ventilation, and air conditioning in the endoscopy unit during non-working hours. We expected this energy consumption to persist even if screening colonoscopies are no longer common. We also ignored the impacts of manufacturing, maintaining, and disposing all test-related equipment. Under counterfactual scenarios, we assumed that hospitals would continue to operate this equipment for other procedures unrelated to primary CRC screening (e.g. diagnostic tests, backup testing for failed/inconclusive primary screening).

Staff transportation was treated as an opportunity cost on sensitivity analyses. This corresponds to FIT-predominant screening scenarios where staff involved with colonoscopies and CTCs redirect their efforts to other activities that do not involve vehicular transportation (e.g. telehealth). We did not account for the transportation of patients undergoing FIT, as completed FITs can be mailed to the laboratory facility. Under sensitivity analyses where staff transportation was considered, we fractionally attributed their impacts over the total number of screening tests that would be conducted by these providers in a typical workday assuming full capacity.

### DATA COLLECTION (Figure 1)

**Figure 1:**
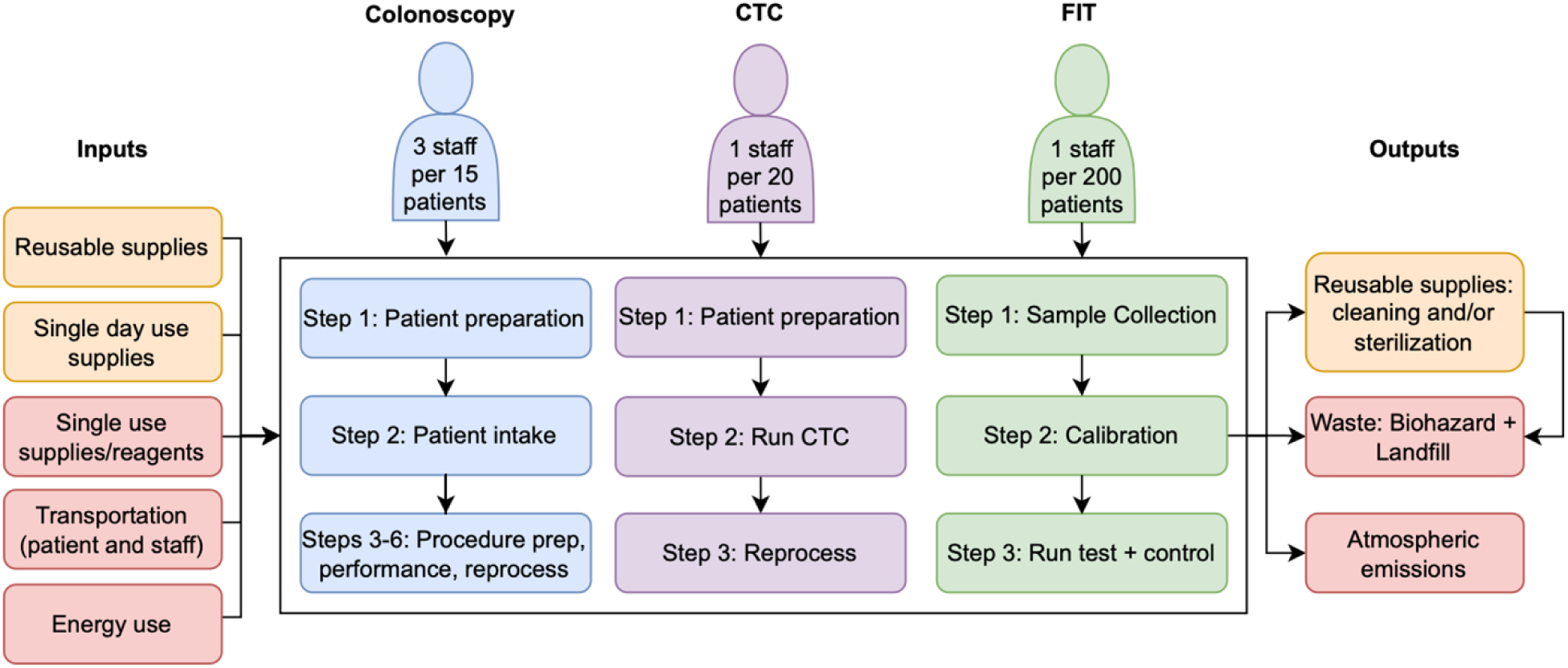
Process map for colorectal cancer screening tests. Blue = colonoscopy process flow, Purple = CTC (CT Colonography) process flow, Green = FIT (Fecal Immunochemical Tests) process flow, Yellow = colonoscopy and CTC inputs and outputs only, Red = inputs and outputs for all 3 processes. For colonoscopy, condensed steps are as follows: step 3: scope retrieval/setup, step 4: anesthesia, step 5: colonoscopy, step 6: scope cleaning/reprocessing. Most of the above steps take place within the hospital, with a few exceptions. Patient bowel preparation (part of step 1 for Colonoscopies and CTCs) and sample collection (FITs) take place at the patient’s home. Transportation occurs to and from the hospital. Procedure outputs (e.g. waste) also occur outside the hospital. All components above correspond to variable (opportunity) costs that are attributable to procedures and thus included in our model. Components not shown and not modelled include the impacts of building and disposing of physical infrastructure, such as the overall hospital facility, colonoscopes, CT scanners, and FIT analyzer. We also did not measure or model any general maintenance activities that are unrelated to the performance of CRC screening tests. Reusable supplies = assumed 75 uses. Single day use supplies = used for multiple procedures in a single day. Single use supplies/reagents = used for single procedure.

Different members of the study team conducted on-site audits on two different days. Their documentation of test procedures were reconciled to minimize the risk of uncaptured elements. Audits involved photo-documentation and identification of all products, manufacturers, and primary material constituents. Masses and/or volumes of all disposable materials were obtained by direct measurement or from the manufacturer’s label. Driving distances for patients and staff were estimated using the Google Maps API.

Energy was measured using plug-load and facility-level energy meters. For CTCs, we performed one phantom scan and five clinical scans to estimate the total attributable energy. Facility-level metering of the endoscopy suite was used to estimate colonoscopy-attributable HVAC utilization. Most endoscopic procedures at UCSF are screening colonoscopies, and nearly all screening colonoscopies at UCSF are performed in this facility. Analogous to our approach for CTCs, we separately measured HVAC energy use during operating hours and after hours and used the difference to estimate colonoscopy-attributable HVAC energy use. These measurements were taken in 3/2023. We did not attribute HVAC-related energy impacts to the FITs or CTCs. These tests represent a small proportion of tests conducted at UCSF and are conducted in rooms shared with other procedures, making it difficult to estimate attributable impacts. Moreover, FIT analyzers are small thus thought unlikely to significantly contribute to HVAC-related impacts.

### DATA MAPPING

Elements were mapped to processes in ecoinvent v3.8 using the allocation at the point of substitution for attribution^10^. Most elements were not identically documented in ecoinvent and thus required additional steps. In most cases, we identified unit processes by reviewing information from the product manufacturer, which often lists the primary material constituent. For some elements not readily identified in ecoinvent, we consulted the LCA literature to identify and use published estimates^11,12^. In other scenarios, we used web search to identify similar products where material constituents were published. We also consulted with an LCA expert on the study team to resolve otherwise unmapped elements, including composite materials. For unit processes that remained uncertain despite the above, we used hierarchical Monte Carlo simulation.

### ESTIMATING IMPACTS OF INDIVIDUAL TESTS

We estimated the impacts of individual screening tests prior to longitudinal modeling. We used ReCiPe2016 weights^13^ to estimate global warming potential [GWP; kg CO_2_ equivalents (kgCO2e)] and damage to human health [DHH; disability adjusted life years (DALYs)]. DHH reflects the total reductions in human health mediated by multiple environmental impacts, including but not limited to global warming. We analyzed the data according to the ‘hierarchist’ perspective, which is based on current scientific consensus on environmental impacts over a 100-year timescale^13^. It does not involve age-weighting or temporal discounting, consistent with DALY estimation methods^14^.

The impacts of car-based transportation were obtained directly from ecoinvent. The impacts of electricity utilization were obtained from ecoinvent using data from the Western Electricity Coordinating Council.

### MARKOV REWARD MODEL

Using these estimates, we constructed a Markov model to estimate the 10-year impacts of each screening strategy (Figure 2). Screening frequencies followed the USPSTF guidelines, with negative FITs repeated every year, negative colonoscopies repeated every 10 years, and negative CTCs repeated every 5 years. Positive FITs and positive CTCs were modeled as resulting in a diagnostic colonoscopy within the same year in simulation time. Our model assumed 100% compliance with all screening procedures. The rationale was that screening compliance varies by geography, and we wanted to obtain results that would hold even with ongoing efforts to achieve 100% screening compliance.

**Figure 2:**
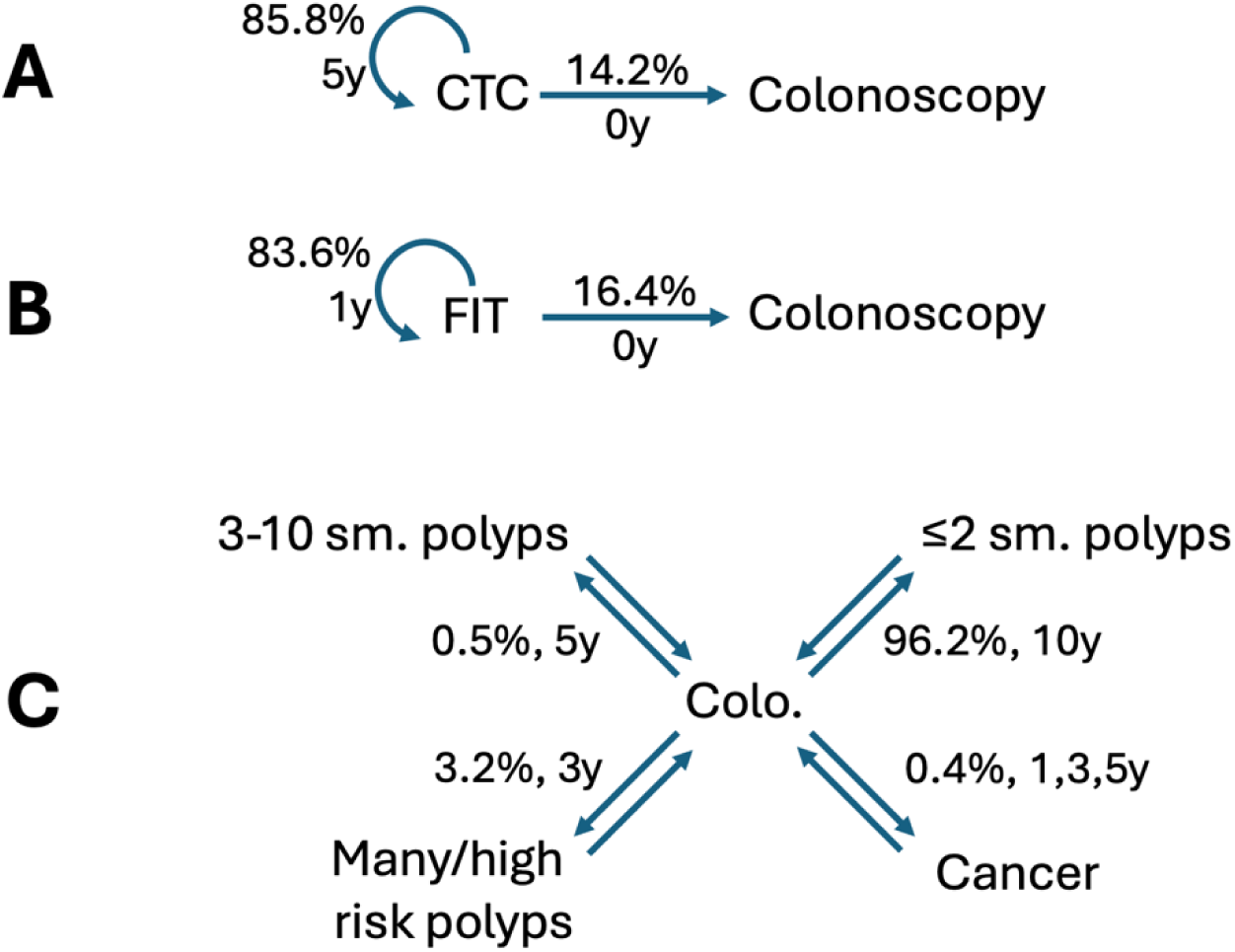
Markov reward model diagram for colorectal cancer screening strategies over a 10-year horizon. Models for CT colonographies, fecal immunochemical tests, and colonoscopies are depicted in panels A, B, and C respectively. Nodes depict states at any given year of the simulation, which are associated with incurring test-level environmental costs. Edges depict transition probabilities and occupancy times, which were parameterized by the literature. CTCs or FITs that yield a positive finding are modeled as resulting in a diagnostic colonoscopy, which itself can yield a variety of findings as shown in panel C.

We used USMSTF guidelines to inform follow-up intervals^15^. Transition probabilities were obtained from the literature^16–18^. In the case of FITs, where positivity rates can vary by test and population, we selected the highest positivity rate in the literature. This choice was informed by our suspicion that FITs would be found superior to the other tests, allowing us to implicitly test the robustness of this conclusion under a “worst case” scenario. We multiplied individual transition probabilities to calculate the probabilities of all possible patient trajectories over 10 years (Supplemental Data).

### STATISTICAL COMPUTING

We used parametric distributions associated with each unit process in ecoinvent to perform hierarchical Monte Carlo simulations. In the first level, we simulated 30 samples from each potentially relevant unit process corresponding to at least one screening strategy. In the second level, we uniformly sampled from each of these distributions and summed component elements to obtain the total impact for each test. We performed 100,000 simulations and calculated the empiric mean and 95% credible intervals for each test. Simulations were performed in Python using the API to openLCA v2.2^19^.

### SENSITIVITY ANALYSES

Our primary analysis was conservative in that it assumes that switching from colonoscopies and CTCs to FITs would result in less vehicular-based transportation by patients only (since their participation involves returning completed tests by mail). Sensitivity analyses relaxed this assumption and assumed that physicians and staff would also no longer travel under a FIT-based regime (e.g. switching to telehealth).

### POLICY MODELING

We modeled the potential consequences of adopting electric vehicles (EVs) on CRC screening impacts. To do this, we replaced the ecoinvent inputs corresponding to car-based transportation with elements corresponding to EVs. To estimate the impacts of transitioning from colonoscopy-predominant screening to FIT-predominant screening, we assumed 1) about 134M patients age-eligible for CRC screening in the US, based on data from the 2020 US census, and 2) 80% current utilization of colonoscopies in the US, contrasted with 80% FIT use in a hypothetical future. To put the potential health savings on an interpretable scale, we converted the DALYs gained per decade to an equivalent gain per year and identified diseases with similar annual morbidity^14^.

## RESULTS

### DATA CAPTURE

We documented 140 unit processes: 33 for FITs, 84 for colonoscopy, and 23 for CTC. These elements mapped to 93 elements within the ecoinvent lifecycle inventory database (Supplemental Tables 1-2, Supplemental Data), as some elements were shared across tests (e.g. gauze, gloves).

### IMPACT OF ISOLATED SCREENING TESTS

We first estimated the impacts of isolated screening tests (Supplemental Table 3, Supplemental Figures 1-2). The attributable emissions of individual FIT tests was 0.27 [0.26, 0.28] kgCO2e.

When accounting for the costs of patient transport to and from UCSF to undergo colonoscopies and CTCs, individual colonoscopies resulted in 39.6 [38.2, 40.5] kgCO2e and CTCs resulted in 27.8 [26.4, 28.7] kgCO2e. When accounting for staff transportation, the impacts of colonoscopies and CTCs increased modestly, to 43.3 [41.6, 44.3] and 28.8 [27.4, 29.8] kgCO2e respectively.

We also estimated the total reductions in human health that are environmentally mediated and attributable to each screening test. These include global warming potential, but also include other effects such as the production of carcinogens, increases in aerosolized particulate matter, ozone depletion, and freshwater consumption ^13^. We found that these environmentally mediated harms were small across tests. Specifically, individual FITs, colonoscopies, and CTCs reduce human health by 0.002, 0.35, and 0.25 DALYs. These effects were robust to the inclusion of staff transport, which affected the results by ∼0.03 DALYs.

The estimated reductions in human health from these tests strongly correlated with their attributable greenhouse gas emissions (0.998 Pearson coefficient). The correlation between these measures remained very strong across all analyses conducted in this study, consistent with public attention to greenhouse gas emissions as the environmental harm of greatest concern.

### LONGITUDINAL IMPACTS

We then compared the 10-year environmental impacts of different screening strategies for average-risk individuals. We assessed three guideline-recommended strategies: 1) annual screening FITs, with positive findings resulting in diagnostic colonoscopies, 2) screening colonoscopies every 10 years, with positive findings resulting in repeat surveillance colonoscopies at potentially sooner intervals, and 3) screening CTCs every 5 years, with positive results resulting in diagnostic colonoscopies^15^.

To estimate longitudinal impacts, we designed a Markov model using parameters informed by the literature. Given the above results that suggested an order-of-magnitude difference favoring FITs over the other tests, we selected the highest FIT positivity rate reported in the literature (16.4%) to test the hypothesized superiority of FITs with maximal robustness^16^. For similar reasons, we assumed that all positive FITs resulted in timely diagnostic colonoscopies, with no delays or lost-to-follow-ups. This allowed us to estimate the upper bound environmental impacts of FIT-first screening, under idealized conditions where target populations are perfectly complaint with health promotion activities.

FIT-first screening was best by a wide margin, resulting in 33.9 [33.6, 34.3] kCO2e of emissions and 0.28 [0.27, 0.29] DALYs lost over 10 years (Table 1; Supplemental Figures 3-4). Colonoscopies were second best, with 41.7 [40.3, 42.7] kCO2e of emissions and 0.34 [0.33, 0.39] DALYs lost. These results were robust to the inclusion of staff-based transportation, which added 3-4 kgCO2e and 0.03-0.06 DALYs of impacts. The significant increase in FIT- and CTC-related impacts were largely attributable to the downstream costs of diagnostic colonoscopies following positive screening tests.

**Table 1:** Estimated impacts of colon cancer screening strategies over 10 years. We report results for two scenarios. The first (“incl. patient transportation”) assumes that staff transportation is not an opportunity cost (e.g. assume that physicians, nurses, technicians and others would still travel from their homes to the medical center to perform other work, under a counterfactual scenario). The second (“incl. transportation by all”) assumes that staff transportation is an opportunity cost. Here, staff transportation is attributed to CTCs and colonoscopies in the sense that they would not travel (e.g. do telehealth instead) under a FIT- predominant scenario. In the latter, the estimates of avoidable impacts (e.g. differences between these estimates and that of FIT as a reference group) are larger in magnitude.

| Screening strategies | Greenhouse gas emissions (kg CO<sub>2</sub> eq.) |  | Damage to Human Health (DALY) |  |
| --- | --- | --- | --- | --- |
|  | Mean | 95% Credible Interval | Mean | 95% Credible Interval |
| FIT | 33.9 | [33.6, 34.3] | 0.28 | [0.27, 0.29] |
| Colo (incl. patient transport) | 41.7 | [40.3, 42.7] | 0.34 | [0.33, 0.39] |
| CTC (incl. patient transport) | 64.2 | [62.9, 65.3] | 0.52 | [0.50, 0.53] |
| Colo (incl. transport by all) | 45.6 | [43.9, 46.7] | 0.37 | [0.36, 0.42] |
| CTC (incl. transport by all) | 67.3 | [65.9, 68.4] | 0.5 | [0.57, 0.60] |

### IMPACT ANALYSIS, POLICY MODELING

We looked for the biggest drivers of impacts by strategy (Table 2). Transportation was the most significant for colonoscopy and CTCs, exceeding the next most significant item by an order of magnitude, and accounting for 44-67% of the total impact of each test. For FITs, transportation’s impacts were similar to paper materials sent by mail.

**Table 2:**
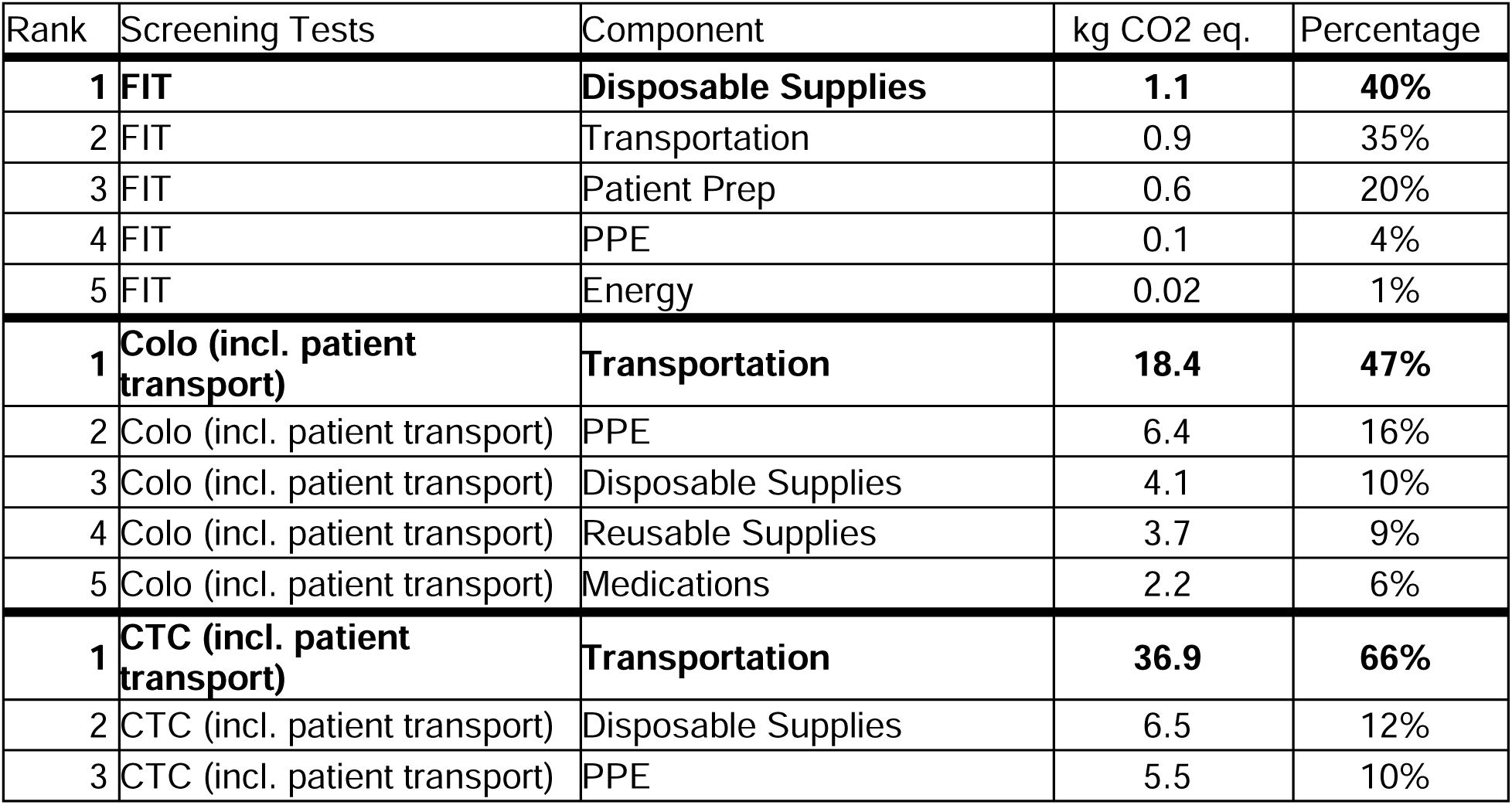
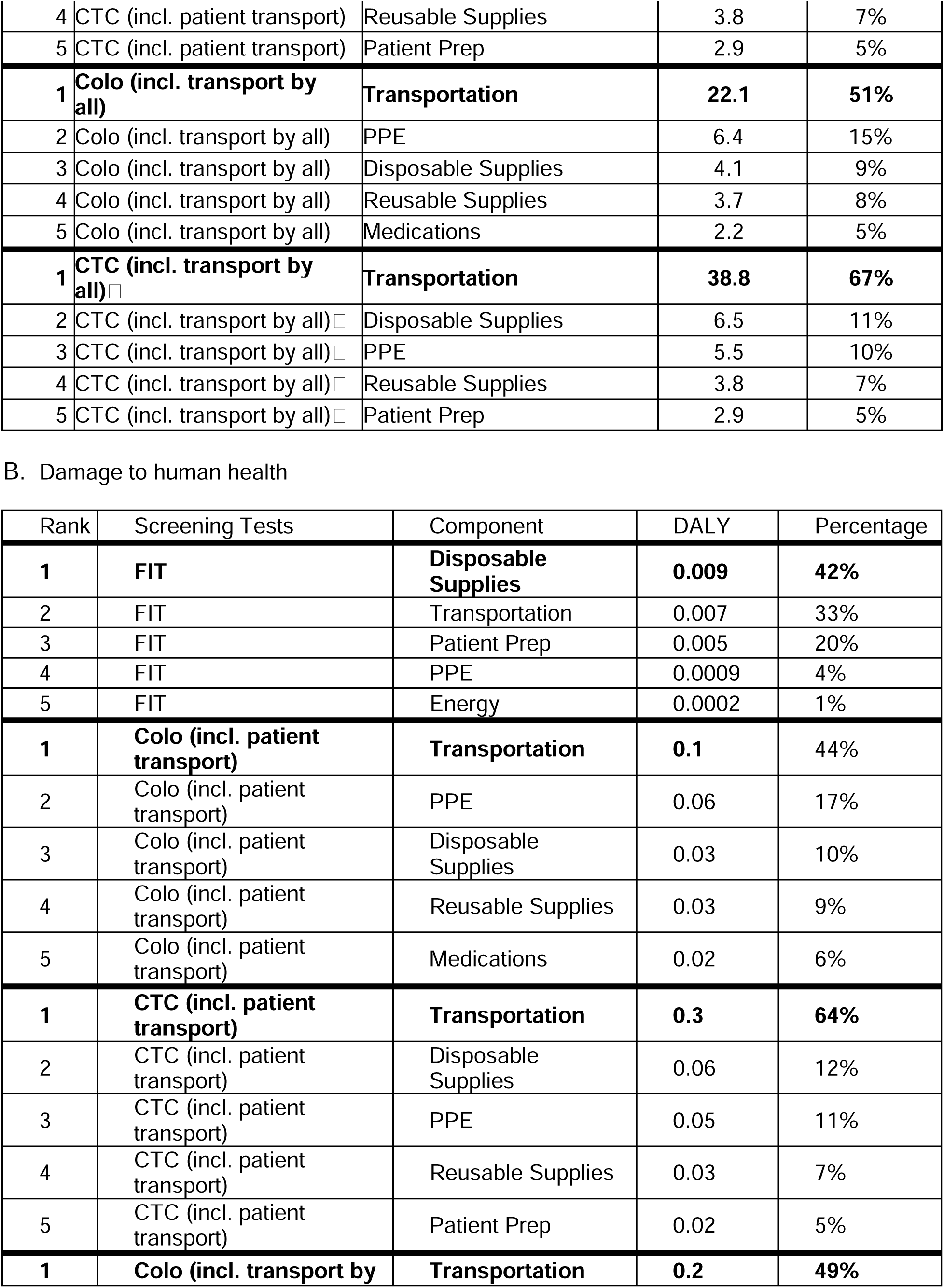
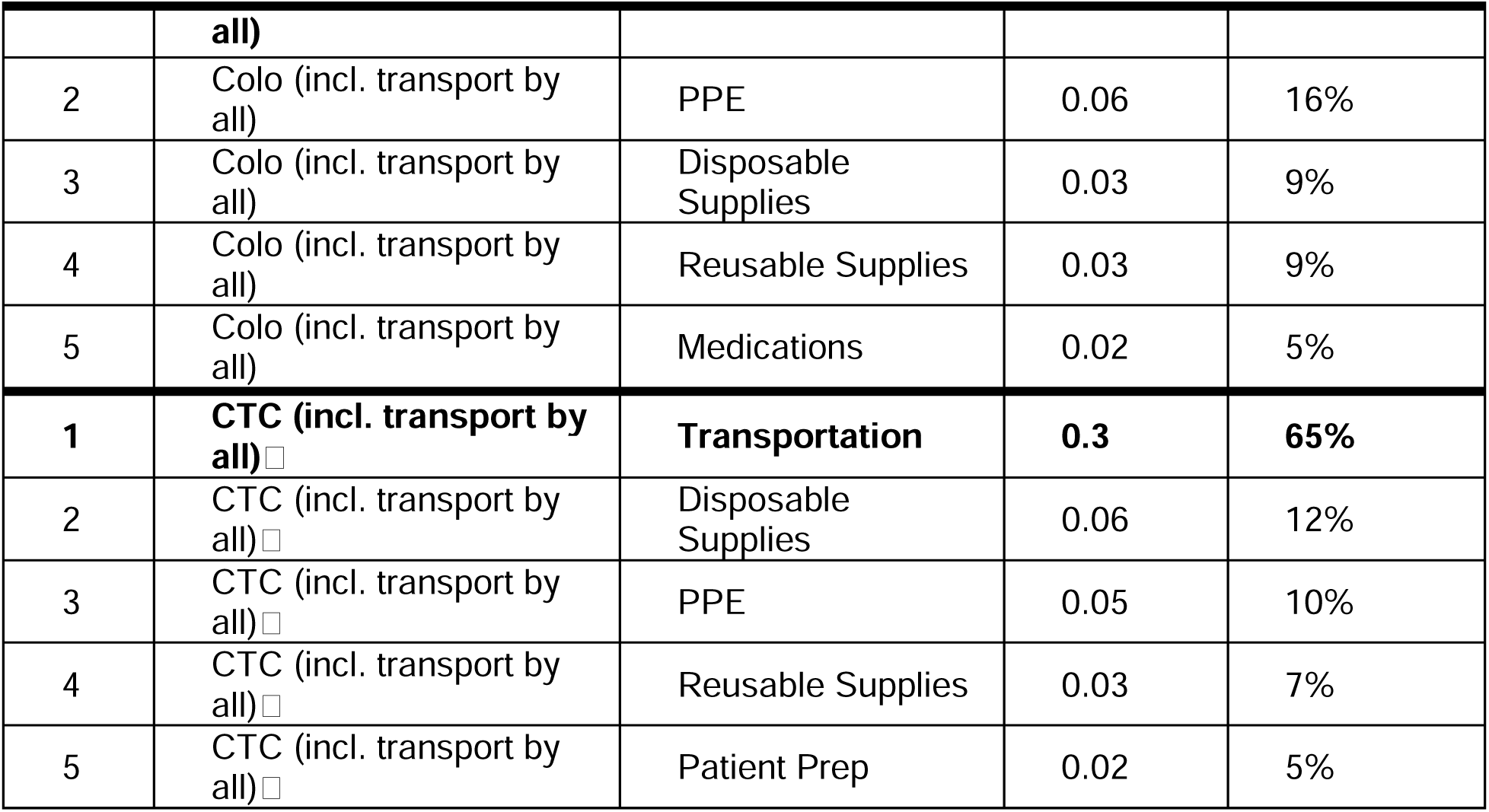
Top five contributors to environmental harms for each screening test. Top contributors within each category are bolded.

A. Greenhouse gas emissions
| Rank | Screening Tests | Component | kg CO<sub>2</sub> eq. | Percentage |
| --- | --- | --- | --- | --- |
| **1** | **FIT** | **Disposable Supplies** | **1.1** | **40%** |
| 2 | FIT | Transportation | 0.9 | 35% |
| 3 | FIT | Patient Prep | 0.6 | 20% |
| 4 | FIT | PPE | 0.1 | 4% |
| 5 | FIT | Energy | 0.02 | 1% |
| **1** | **Colo (incl. patient transport)** | **Transportation** | **18.4** | **47%** |
| 2 | Colo (incl. patient transport) | PPE | 6.4 | 16% |
| 3 | Colo (incl. patient transport) | Disposable Supplies | 4.1 | 10% |
| 4 | Colo (incl. patient transport) | Reusable Supplies | 3.7 | 9% |
| 5 | Colo (incl. patient transport) | Medications | 2.2 | 6% |
| **1** | **CTC (incl. patient transport)** | **Transportation** | **36.9** | **66%** |
| 2 | CTC (incl. patient transport) | Disposable Supplies | 6.5 | 12% |
| 3 | CTC (incl. patient transport) | PPE | 5.5 | 10% |
| 4 | CTC (incl. patient transport) | Reusable Supplies | 3.8 | 7% |
| 5 | CTC (incl. patient transport) | Patient Prep | 2.9 | 5% |
| **1** | **Colo (incl. transport by all)** | **Transportation** | **22.1** | **51%** |
| 2 | Colo (incl. transport by all) | PPE | 6.4 | 15% |
| 3 | Colo (incl. transport by all) | Disposable Supplies | 4.1 | 9% |
| 4 | Colo (incl. transport by all) | Reusable Supplies | 3.7 | 8% |
| 5 | Colo (incl. transport by all) | Medications | 2.2 | 5% |
| **1** | **CTC (incl. transport by all)□** | **Transportation** | **38.8** | **67%** |
| 2 | CTC (incl. transport by all)□ | Disposable Supplies | 6.5 | 11% |
| 3 | CTC (incl. transport by all)□ | PPE | 5.5 | 10% |
| 4 | CTC (incl. transport by all)□ | Reusable Supplies | 3.8 | 7% |
| 5 | CTC (incl. transport by all)□ | Patient Prep | 2.9 | 5% |

| Rank | Screening Tests | Component | DALY | Percentage |
| --- | --- | --- | --- | --- |
| **1** | **FIT** | **Disposable Supplies** | **0.009** | **42%** |
| 2 | FIT | Transportation | 0.007 | 33% |
| 3 | FIT | Patient Prep | 0.005 | 20% |
| 4 | FIT | PPE | 0.0009 | 4% |
| 5 | FIT | Energy | 0.0002 | 1% |
| **1** | **Colo (incl. patient transport)** | **Transportation** | **0.1** | **44%** |
| 2 | Colo (incl. patient transport) | PPE | 0.06 | 17% |
| 3 | Colo (incl. patient transport) | Disposable Supplies | 0.03 | 10% |
| 4 | Colo (incl. patient transport) | Reusable Supplies | 0.03 | 9% |
| 5 | Colo (incl. patient transport) | Medications | 0.02 | 6% |
| **1** | **CTC (incl. patient transport)** | **Transportation** | **0.3** | **64%** |
| 2 | CTC (incl. patient transport) | Disposable Supplies | 0.06 | 12% |
| 3 | CTC (incl. patient transport) | PPE | 0.05 | 11% |
| 4 | CTC (incl. patient transport) | Reusable Supplies | 0.03 | 7% |
| 5 | CTC (incl. patient transport) | Patient Prep | 0.02 | 5% |
| **1** | **Colo (incl. transport by** **all)** | **Transportation** | **0.2** | **49%** |
| 2 | Colo (incl. transport by all) | PPE | 0.06 | 16% |
| 3 | Colo (incl. transport by all) | Disposable Supplies | 0.03 | 9% |
| 4 | Colo (incl. transport by all) | Reusable Supplies | 0.03 | 9% |
| 5 | Colo (incl. transport by all) | Medications | 0.02 | 5% |
| **1** | **CTC (incl. transport by all)**□ | **Transportation** | **0.3** | **65%** |
| 2 | CTC (incl. transport by all)□ | Disposable Supplies | 0.06 | 12% |
| 3 | CTC (incl. transport by all)□ | PPE | 0.05 | 10% |
| 4 | CTC (incl. transport by all)□ | Reusable Supplies | 0.03 | 7% |
| 5 | CTC (incl. transport by all)□ | Patient Prep | 0.02 | 5% |

These transportation-related impacts, as estimated using ecoinvent, implicitly utilize an average of car engine types (internal combustion, electric) and vehicle sizes representative of the global passenger car market^10^. As such, our results reflect the current predominance of gasoline-based transportation.

We explored the potential impacts of recent policies to decarbonize transportation. For instance, in 2022 the California Air Resources Board approved a roadmap that requires new vehicles sold in 2035 to be zero-emission^20^. Accordingly, we used ecoinvent data to simulate the impacts of transitioning from 0% to 50% EV use. This would result in 7-10% reductions in kCO2e, and 5-7% reductions in DALYs lost (Table 3; Supplemental Table 4).

**Table 3:** Reductions in environmental impacts following increasing adoption of electric vehicles. Results are derived from the calculations reported in supplemental table 4. Colonoscopy and CTC results correspond to the scenario where only patient transportation is modeled (provider and staff transportation are treated as fixed/unavoidable costs and thus not included in the model).

|  |  | Greenhouse gas emissions (kg CO<sub>2</sub> eq.) |  | Damage to Human Health (DALY) |  |
| --- | --- | --- | --- | --- | --- |
| Screening strategies | EV change in utilization | Absolute reduction | Relative reduction | Absolute reduction | Relative reduction |
| FIT | 0%-50% | 2.5 | 7% | 0.02 | 6% |
| FIT | 0%-100% | 5.0 | 15% | 0.03 | 11% |
| Colonoscopy | 0%-50% | 3.1 | 8% | 0.02 | 6% |
| Colonoscopy | 0%-100% | 6.3 | 15% | 0.04 | 11% |
| CTC | 0%-50% | 6.4 | 10% | 0.04 | 8% |
| CTC | 0%-100% | 12.9 | 20% | 0.08 | 15% |

Finally, we estimated the impact of transitioning to FIT-predominant screening in the US, a country where colonoscopies predominate. Assuming ∼134M patients eligible for CRC screening annually, and assuming that the current utilization of screening colonoscopies is about 80% (with the remainder being FIT or FIT-like tests), transitioning to 80% FIT-based screening would result in 5.2 million disability adjusted life years gained every 10 years. To put this in perspective, the potential benefits to global health are roughly equivalent to curing and preventing all worldwide cases of cleft lip/palate, or nephrolithiasis, or testicular cancer^14^.

## DISCUSSION

This is the first study to compare the environmental impacts of three guideline-recommended screening strategies for CRC: FITs, colonoscopies, and CTCs. We found that FITs were the most environmentally sustainable screening strategy, roughly 20% better than colonoscopies. In our analysis, CTCs came out least favorably. The largest driver of harms for colonoscopies and CTC was gasoline-predominant transportation. This finding largely explains why FITs came out as best: they are the only tests that can be conducted by patients without travel to a healthcare facility. FITs as isolated tests were over 100 times superior to colonoscopies and CTCs. This degree of superiority was curbed in the Markov model, after accounting for downstream colonoscopies following positive FITs. Nonetheless, even under a worst-case modeling scenario that incorporates the highest rates of FIT positivity in the literature, FIT-first screening was able to prevent enough downstream colonoscopies to remain superior.

### COMPARISON TO PREVIOUS LITERATURE

The most relevant prior study on this topic was conducted by Desai et al^21^. The authors audited gastrointestinal endoscopy procedures at their center, focusing on physical waste and energy utilization. They estimated the average carbon footprint of a GI procedure to be 1.5 kgCO2e, which contrasts markedly with our estimate of 39.6 kgCO2e per colonoscopy. We attribute this to differences in study goals and estimation methods. Desai et al. sought to study and inform practices within the endoscopy suite, whereas we sought to study and inform CRC screening procedures. As such, we included patient and staff transportation as an opportunity cost, whereas Desai et al did not. Transportation accounted for about half of the environmental impact from colonoscopies, explaining the order of magnitude difference in our estimates. Another study of endoscopic practices in France found that that transportation was indeed the largest single driver of greenhouse gas emissions, consistent with our findings^22^.

### POLICY IMPLICATIONS

Prioritizing FITs over colonoscopies across the US could lead to substantial gains for global health while also reducing the costs of care. We found that the benefits to global health are comparable to curing and preventing all global cases of cleft lip/palate, or nephrolithiasis, or testicular cancer. From a screening effectiveness standpoint, both tests are recommended by the USPSTF and are thought similarly effective and safe. However, recent cost-effectiveness studies favor FITs over colonoscopies, with an incremental cost-effectiveness ratio of nearly $1M per quality-adjusted life year gained by colonoscopies compared to annual FIT-based screening^23^.

Our findings call for urgent action by payors, who strongly influence what services occur. The case for FIT-based screening should be clear, given the alignment between its cost-effectiveness and environmental harm reduction. Related to this point, we believe that health technology assessments need to embrace a broader view of cost-benefit analyses that include environmental harms to future generations^24,25^. Public health authorities, medical guideline authors, and physicians also have a role to play in making this policy proposal a reality.

Although our study directly speaks to the environmental impacts of CRC screening strategies, it has indirect implications on other ways that health systems and governments can promote more sustainable practices. One example is telehealth. To the extent that remote visits can promote patient health just as well as in person visits in many cases, payors and clinics should continue this practice that avoids unnecessary driving by patients and providers.

Governments can promote sustainability by incentivizing environmental product declarations across the economy. About 80% of the attributable emissions from healthcare activities are indirect (scope 3) and occur in the supply chain^2^. Although these emissions are not under an organization’s direct control, they can influence these effects by choosing vendors with lower emissions.

Unfortunately, this kind of data is currently not available to consumers. Fortunately, recent policies like the EU’s Corporate Sustainability Reporting Directive (CSRD) and the US EPA’s Greenhouse Gas Reporting Program that require aggregate reporting of emissions data are now changing this. Extending these frameworks to the product level and making this information accessible to institutional consumers are key next steps that governments can facilitate.

Our analysis suggests that transitioning to electric vehicles (EVs) could yield modest additional benefits, and governments have a key role in facilitating this. Specifically, our analysis suggests that society-level transitions to all EVs in the San Francisco area could reduce CRC-attributable emissions by 15-20%. Continued innovations in battery manufacturing and greater utilization of renewable energy in the electrical grid may further enhance the benefits of transitioning to EVs.

### STRENGTHS, LIMITATIONS

This study builds on prior work using more robust methods. In contrast to the study by Desai et al. this study incorporates LCAs, the gold-standard for estimating environmental impacts. We used ecoinvent, the most comprehensive life cycle inventory, to inform our estimates and to conduct probabilistic sensitivity analyses. This study is, to our knowledge, the first to integrate LCAs with Markov Reward Modeling to make longitudinal assessments. We used this approach to account for the potential outcomes of different screening strategies, incorporating worst-case parameters for FIT model for added robustness. Lastly, to make our study more accessible to healthcare audiences, we also report impacts by DALYs lost.

This study has some limitations. For one, this is a single center assessment with a US focus. It is likely that there are variations in how procedures are conducted across centers and countries. We identified transportation as the dominant driver of environmental harms, and these impacts will vary by geography. Transportation-related impacts will likely be worse in rural areas with less connectivity, and better in cities with public transportation. We think these effects are unlikely to affect our finding of FIT’s superiority, but additional studies are needed. Second, our study relied on the accuracy of ecoinvent, which has limited data on pharmaceutical manufacturing and distribution. This is due to the current lack of incentives for environmental product reporting. Hopefully this will change as laws like CSDR take hold. As before, given the greater use of medications in colonoscopies and CTCs compared to FITs, we expect that the lack of reliable data here will not affect our main conclusions. Third, this study only investigated three screening strategies. Future studies will be needed to incorporate additional strategies, including those using blood-based biomarker tests. Lastly, our models incorporate many implicit and difficult to test assumptions, particularly about counterfactual scenarios. For example, staff transportation if FITs are prioritized as a screening test. More studies exploring a broader range of modeling scenarios are needed.

## CONCLUSION

FITs are more environmentally sustainable for CRC screening than colonoscopies or CT Colonographies. Given the similar efficacy and safety of these tests, payors and clinicians should favor FITs for those at low risk for colorectal cancer. Efforts to decarbonize transportation and electric grids, incentivize telehealth, and mandate environmental product declarations and sustainable product design will help curb the unintended harms of healthcare, and promote the health of current and future generations worldwide.

## Supporting information

Supplemental Results

Data Worksheet

Sample Data

Description of Data Inputs

## ACKNOWLEDGEMENTS

The authors thank the staff of the UCSF gastroenterology, radiology, and laboratory medicine departments for facilitating on-site audits and answering technical questions about all CRC screening procedures.

## DATA AVAILABILITY

An analysis-ready dataset and analytical code accompany this manuscript as supplemental files.

## AUTHOR CONTRIBUTIONS

VAR conceived the study. VAR and SG obtained funding. TW, KW, NA, PV, GM, and AB collected the data. TW processed and analysed the data. SW, WK, SP, CLT, SG, SW, and VAR provided oversight. VAR wrote the first draft of the manuscript. All authors interpreted the data and critically edited the manuscript.

## REFERENCES

1. Climate change and noncommunicable diseases: connections. Accessed August 31, 2024. https://www.who.int/news/item/02-11-2023-climate-change-and-noncommunicable-diseases-connections

2. Eckelman MJ, Huang K, Lagasse R, Senay E, Dubrow R, Sherman JD. Health Care Pollution And Public Health Damage In The United States: An Update. Health Aff (Millwood). 2020;39(12):2071–2079. doi:10.1377/hlthaff.2020.01247

3. Tennison I, Roschnik S, Ashby B, et al. Health care’s response to climate change: a carbon footprint assessment of the NHS in England. Lancet Planet Health. 2021;5(2):e84–e92. doi:10.1016/S2542-5196(20)30271-0

4. US Preventive Services Task Force. Screening for Colorectal Cancer: US Preventive Services Task Force Recommendation Statement. JAMA. 2021;325(19):1965–1977. doi:10.1001/jama.2021.6238

5. Lin JS, Perdue LA, Henrikson NB, Bean SI, Blasi PR. Screening for Colorectal Cancer: Updated Evidence Report and Systematic Review for the US Preventive Services Task Force. JAMA. 2021;325(19):1978–1998. doi:10.1001/jama.2021.4417

6. Research iData. An Astounding 16.6 Million Colonoscopies are Performed Annually in The United States. iData Research. August 8, 2018. Accessed August 31, 2024. https://idataresearch.com/an-astounding-19-million-colonoscopies-are-performed-annually-in-the-united-states/

7. PAHO/WHO. Colorectal Cancer Screening in the Americas. https://www3.paho.org/hq/dmdocuments/2016/Colorectal-Cancer-Screening-Landscape-English.pdf

8. United European Gastroenterology. COLORECTAL SCREENING ACROSS EUROPE. https://ueg.eu/files/779/67d96d458abdef21792e6d8e590244e7.pdf

9. Li J, Yao H, Lu Y, Zhang S, Zhang Z, Society of Digestive Endoscopy of the Chinese Medical Association, Colorectal Surgery Group of the Chinese Medical Association, Chinese Association of Gastroenterologist & Hepatologist, National Clinical Research Center for Digestive Diseases, Chinese Medical Journal Clinical Practie Guideline Collaborative. Chinese national clinical practice guidelines on prevention, diagnosis and treatment of early colorectal cancer. Chin Med J (Engl). 2024;137(17):2017–2039. doi:10.1097/CM9.0000000000003253

10. Wernet G, Bauer C, Steubing B, Reinhard J, Moreno-Ruiz E, Weidema B. The ecoinvent database version 3 (part I): overview and methodology. Int J Life Cycle Assess. 2016;21(9):1218–1230. doi:10.1007/s11367-016-1087-8

11. Ponder CS. Life Cycle Inventory Analysis of Medical Textiles and Their Role in Prevention of Nosocomial Infections. Published online December 7, 2009. Accessed August 31, 2024. http://www.lib.ncsu.edu/resolver/1840.16/4715

12. Thiel CL, Sreedhar P, Silva GS, et al. Effectiveness of Conservation Practices for Personal Protective Equipment: A Systematic Review. *Available SSRN 4071361.* Published online 2022. Accessed August 31, 2024. https://papers.ssrn.com/sol3/papers.cfm?abstract_id=4071361

13. Huijbregts MAJ, Steinmann ZJN, Elshout PMF, et al. ReCiPe2016: a harmonised life cycle impact assessment method at midpoint and endpoint level. Int J Life Cycle Assess. 2017;22(2):138–147. doi:10.1007/s11367-016-1246-y

14. Global Health Estimates. Accessed August 31, 2024. https://www.who.int/data/global-health-estimates

15. Gupta S, Lieberman D, Anderson JC, et al. Recommendations for Follow-Up After Colonoscopy and Polypectomy: A Consensus Update by the US Multi-Society Task Force on Colorectal Cancer. Gastrointest Endosc. 2020;91(3):463–485.e5. doi:10.1016/j.gie.2020.01.014

16. Alsayid M, Singh MH, Issaka R, et al. Yield of Colonoscopy After a Positive Result From a Fecal Immunochemical Test OC-Light. Clin Gastroenterol Hepatol Off Clin Pract J Am Gastroenterol Assoc. 2018;16(10):1593–1597.e1. doi:10.1016/j.cgh.2018.04.014

17. Yee J. Screening CT Colonography Offers Improved Diagnostic Performance Compared with Multitarget Stool DNA Testing. Radiology. 2020;297(1):130–131. doi:10.1148/radiol.2020202856

18. Regula J, Rupinski M, Kraszewska E, et al. Colonoscopy in Colorectal-Cancer Screening for Detection of Advanced Neoplasia. N Engl J Med. 2006;355(18):1863–1872. doi:10.1056/NEJMoa054967

19. Ciroth A. ICT for environment in life cycle applications openLCA — A new open source software for life cycle assessment. Int J Life Cycle Assess. 2007;12(4):209–210. doi:10.1065/lca2007.06.337

20. California moves to accelerate to 100% new zero-emission vehicle sales by 2035 | California Air Resources Board. Accessed August 31, 2024. https://ww2.arb.ca.gov/news/california-moves-accelerate-100-new-zero-emission-vehicle-sales-2035

21. Desai M, Campbell C, Perisetti A, et al. The Environmental Impact of Gastrointestinal Procedures: A Prospective Study of Waste Generation, Energy Consumption, and Auditing in an Endoscopy Unit. Gastroenterology. 2024;166(3):496–502.e3. doi:10.1053/j.gastro.2023.12.006

22. Lacroute J, Marcantoni J, Petitot S, et al. The carbon footprint of ambulatory gastrointestinal endoscopy. Endoscopy. 2023;55:918–926. doi:10.1055/a-2088-4062

23. Ladabaum U, Mannalithara A, Weng Y, et al. Comparative Effectiveness and Cost-Effectiveness of Colorectal Cancer Screening With Blood-Based Biomarkers (Liquid Biopsy) vs Fecal Tests or Colonoscopy. Gastroenterology. 2024;167(2):378–391. doi:10.1053/j.gastro.2024.03.011

24. Navigating the Intersection of Healthcare and Environmental Sustainability: ISPOR Europe round-up on the inclusion of environmental impact in HTA - OHE. OHE - Leading intellectual authority on global health economics. January 17, 2024. Accessed August 31, 2024. https://www.ohe.org/insight/ispor-healthcare-environmental-sustainability

25. Williams JTW, Bell KJL, Morton RL, Dieng M. Methods to Include Environmental Impacts in Health Economic Evaluations and Health Technology Assessments: A Scoping Review. Value Health. 2024;27(6):794–804. doi:10.1016/j.jval.2024.02.019

