## Supplemental Methods for "A Comparative Environmental Impact Analysis of Screening Tests for Colorectal Cancer"

DATA COLLECTION

At UCSF, screening colonoscopies are usually performed using meperidine, midazolam, and ondansetron for sedation; these were included in our study. We did not capture data on materials used to perform biopsies or polypectomies (e.g. cold snares), which might be conducted during a colonoscopy. These materials are uncommonly used in average risk patients aged 45, and were thought *a priori* to be negligible in their environmental impacts relative to other component processes given work by Gordon et al.

Gordon IO, Sherman JD, Leapman M, Overcash M, Thiel CL. Life Cycle Greenhouse Gas Emissions of Gastrointestinal Biopsies in a Surgical Pathology Laboratory. *Am J Clin Pathol*. 2021;156(4):540-549. doi:10.1093/ajcp/aqab021

CT COLONOGRAPHY PROCESS MAP

A process map was generated to define the steps and associated time intervals involved with CT colonography exam. This was done through prospective observations of the CT colonography exam by individuals trained to observe environmental impact. The observations were made at different imaging locations, CT scanners, and with different performing technologists to observe potential variations that might exist in our CT workflow. Patients were also interviewed to determine activities performed before arriving at the hospital to inform prearrival impact.

CT COLONOGRAPHY ENERGY ESTIMATION

*CT unit*

The CT unit included a representative scanner that the majority of CT colonography exams are performed. The CT unit was equipped with a power meter (SEM3 or Sentron PAC4200 Power Meters, Siemens Smart Infrastructure). The CT was a 64 slice LightSpeed VCT (General Electric) installed in 2008.

*Power Measurements*

Power meter locations were selected by an electrical engineer to isolate native CT functions. Power meters were installed in individual equipment rooms at the main electrical panel serving the CT suite and provided continuous monitoring with a 1-Hz sampling rate. Energy data were sent by local area networks to the building management system (Disego CC; Siemens).

Power measurement logs were extracted on days of five different CT colonography exams. Power data were recorded in kilowatts with a temporal resolution of 10 seconds. Data for each scanner was segmented into individual scans defined as the time period between the start of the localizer acquisition to the end of the last scan. Total energy and time were calculated for each exam. Data was reported with descriptive statistics for modeling experiments.
