## Supplemental Results for "A Comparative Environmental Impact Analysis of Screening Tests for Colorectal Cancer"

**Table of Contents**

**Supplemental Table 1: Input elements collected by onsite audits.**

**Supplemental Table 2: Modeling elements used in the life cycle inventory database**

**Supplemental Table 3: Estimated impacts of individual colon cancer screening tests**

**Supplemental Table 4: Mean reductions in environmental impacts following transitions to electric vehicles**

**Supplemental Figure 1: Estimated greenhouse gas emissions resulting from performing individual CRC screening tests**

**Supplemental Figure 2: Estimated damage to human health resulting from performing individual CRC screening tests**

**Supplemental Figure 3: Projected greenhouse gas emissions resulting from different screening strategies over 10 years**

**Supplemental Figure 4: Projected damage to human health resulting from different screening strategies over 10 years**

**Supplemental Table 1: Input elements collected by onsite audits.** All 140 inputs manually documented during onsite audits of testing sites are reported here.

| Input Names | | |
| --- | --- | --- |
| 1ml Negative Control Vial (with negative control inside) | Sterile Water 500ml - Plastic | IV Starter kit |
| laundry electricity for patient gown, pillow and blanket | Channel check strip - paper | Shoe covers |
| 1ml Positive Control Vial (with positive control inside) | ChloraPrep Single Swabstick | Mailing Envelope |
| First step Endoscope Bedside Pre-clean Kit (200ml) | Cuvette Box Packaging | Red Bin Cover |
| Dri Scope Aid Disposable Tubing Kit packaging | Valve and Connector Kit | 1ml of CLRW |
| laundry water for patient gown, pillow and blanket | Disinfection wipes | Heated blanket |
| Dri Scope Aid Disposable Tubing Kit | 30cc Syringe water | Latex Reagent |
| Dri Scope Aid Disposable Connectors Kit | Wash Concentrate | Oxygen Tube |
| 485*4ml CLRW for Wash Concentrate | Oxygen Tube packaging | Chux pad |
| Protocol Administration set - 35 mL syringe | IV bag LR 1L - Chemical | IV Catheter |
| Latex Reagent Bottle (without 4ml reagent) | Surgical lubricant - gel | IV tubing |
| endoscope cleaning brush packaging (yellow) | Surgical mask (Type II) | CO2 Tubing |
| 4mg medication vial for ondansetron | Sterile Water 1L - Plastic | Face shield |
| Protocol Administration set - flexible tubing | Energy for reprocessing | Thermal Paper |
| Main Packaging and other papers | double-end brush (green) | Packet of Gauze (5) |
| Latex Packaging (with papers for 2 vials) | Sheet*2 (Fitted + draw) | Patient gown |
| endoscope cleaning brush (yellow) | 45 ml Diluent Bottle | Pillow Case |
| Calibration Kit Packaging (10 per pack) | IV bag LR 1L - Water | 3 ml Syringe |
| Negative Control Packaging (5 per pack) | Surgical lubricant - Plastic | Cuvettes |
| 100mg medication vial for meperidine | Tergal Detergent | Buffer Media |
| 5mg medication vial for midazolam | Sterile Water 1L - Water | Buffer Packaging |
| Anesthesia glass vials for all 3 (1,2,5mL) | Plastic sheet for scope | Green Bin Cover |
| Green Red/Bin/liner Cover packaging | Suction Tubing packaging | Patient Robe |
| 50cc Syringe without sterile water | 1 cidex test strip/run | Pillow cover |
| Positive Control Packaging (5 per pack) | Blood pressure cuff | 5 ml Syringe |
| Box of Cuvettes (without cuvettes) | Plastic Bin Liner | GOLYTELY |
| Channel check strip - foil packaging | Suction Tubing | Parafilm |
| Suprep Bowel Prep Kit - medication bottle*2 | Suction Canister (empty) | Towel |
| Suprep Bowel Prep Kit - one cup | Total Endoscopy energy | renuzyme |
| Valve and Connector Kit packaging | transportation km | Bouffant |
| Suprep Bowel Prep Kit - chemical*2 | Total FIT energy | Bed sheet |
| Suprep Bowel Prep Kit - tap water | Triangle cannister | Gloves |
| Chux pad Ultrasorbs Advanced 350 | Tube gel - container | Socks |
| Sterile Water 500ml - Water | IV tubing packaging | Media |
| Set of telemetry strips (EKG) | Irrigation Tubing | Cidex |
| Wash Concentrate Packaging | CTC total energy | Gown |
| Oxygen tube tip packaging | 1ml of Calibrator | Cup |
| Sampling Bottle (without media) | Tube gel - gel |  |

**Supplemental Table 2: Modeling elements used in the life cycle inventory database.** All 93 LCA inputs, using the nomenclature associated with the ecoinvent v3.8 APOS database, are listed here. GLO = Global (averaged over all countries in the ecoinvent database). RoW = Rest of World. In ecoinvent, this typically refers to countries outside of Europe.

| Input Name | Area | Input Name | Area |
| --- | --- | --- | --- |
| market for polyethylene terephthalate, granulate, amorphous, recycled \| polyethylene terephthalate, granulate, amorphous, recycled \| APOS, U | - US | market for glass fibre reinforced plastic, polyester resin, hand lay-up \| glass fibre reinforced plastic, polyester resin, hand lay-up \| APOS, U | - GLO |
| market for sodium hypochlorite, without water, in 15% solution state \| sodium hypochlorite, without water, in 15% solution state \| APOS, U | - RoW | market for extrusion of plastic sheets and thermoforming, inline \| extrusion of plastic sheets and thermoforming, inline \| APOS, U | - GLO |
| market for glass cullet, mixed glass from used cathode ray tube \| glass cullet, mixed glass from used cathode ray tube \| APOS, U | - GLO | market for carbon fibre reinforced plastic, injection moulded \| carbon fibre reinforced plastic, injection moulded \| APOS, U | - GLO |
| market for carton board box production, with gravure printing \| carton board box production, with gravure printing \| APOS, U | - GLO | market for polystyrene foam slab with graphite, 6% recycled \| polystyrene foam slab with graphite, 6% recycled \| APOS, U | - GLO |
| market for polyurethane, flexible foam, flame retardant \| polyurethane, flexible foam, flame retardant \| APOS, U | - GLO | market for solid bleached and unbleached board carton \| solid bleached and unbleached board carton \| APOS, U | - RoW |
| market for polyethylene, linear low density, granulate \| polyethylene, linear low density, granulate \| APOS, U | - GLO | market for paper, woodcontaining, lightweight coated \| paper, woodcontaining, lightweight coated \| APOS, U | - RoW |
| market for urea formaldehyde foam, in situ foaming \| urea formaldehyde foam, in situ foaming \| APOS, U | - GLO | market for aluminium collector foil, for Li-ion battery \| aluminium collector foil, for Li-ion battery \| APOS, U | - GLO |
| market for packaging film, low density polyethylene \| packaging film, low density polyethylene \| APOS, U | - GLO | market for funnel glass, for cathode ray tube display \| funnel glass, for cathode ray tube display \| APOS, U | - GLO |
| market for polyethylene, high density, granulate \| polyethylene, high density, granulate \| APOS, U | - GLO | market for polystyrene foam slab, 10% recycled \| polystyrene foam slab, 10% recycled \| APOS, U | - GLO |
| market for packaging, for fertilisers or pesticides \| packaging, for fertilisers or pesticides \| APOS, U | - GLO | market for dipropylene glycol monomethyl ether \| dipropylene glycol monomethyl ether \| APOS, U | - RoW |
| market for carbon dioxide, in chemical industry \| carbon dioxide, in chemical industry \| APOS, U | - GLO | market for waste polyethylene terephthalate \| waste polyethylene terephthalate \| APOS, U | - RoW |
| market for transport, passenger car, electric \| transport, passenger car, electric \| APOS, U | - GLO | market for hazardous waste, for incineration \| hazardous waste, for incineration \| APOS, U | - RoW |
| market for polyvinylchloride, bulk polymerised \| polyvinylchloride, bulk polymerised \| APOS, U | - GLO | market for textile, nonwoven polypropylene \| textile, nonwoven polypropylene \| APOS, U | - GLO |
| market for liquid packaging board container \| liquid packaging board container \| APOS, U | - GLO | market for white lined chipboard carton \| white lined chipboard carton \| APOS, U | - RoW |
| market for containerboard, fluting medium \| containerboard, fluting medium \| APOS, U | - CA- QC | market for polyurethane, flexible foam \| polyurethane, flexible foam \| APOS, U | - RoW |
| market for containerboard, linerboard \| containerboard, linerboard \| APOS, U | - CA- QC | market for textile, nonwoven polyester \| textile, nonwoven polyester \| APOS, U | - GLO |
| market for sulfate pulp, bleached \| sulfate pulp, bleached \| APOS, U | - RoW | market for corrugated board box \| corrugated board box \| APOS, U | - RoW |
| market for thermo-mechanical pulp \| thermo-mechanical pulp \| APOS, U | - RoW | market for paper, woodfree, uncoated \| paper, woodfree, uncoated \| APOS, U | - RoW |
| market for electricity, high voltage \| electricity, high voltage \| APOS, U | - US-WECC | market for sulfate pulp, unbleached \| sulfate pulp, unbleached \| APOS, U | - RoW |
| market for electricity, medium voltage \| electricity, medium voltage \| APOS, U | - US-WECC | market for paper, woodfree, coated \| paper, woodfree, coated \| APOS, U | - RoW |
| market for electricity, low voltage \| electricity, low voltage \| APOS, U | - US-WECC | market for aluminium alloy, AlMg3 \| aluminium alloy, AlMg3 \| APOS, U | - GLO |
| market for ethylvinylacetate, foil \| ethylvinylacetate, foil \| APOS, U | - GLO | market for packaging, for pesticides \| packaging, for pesticides \| APOS, U | - GLO |
| market for extrusion, plastic pipes \| extrusion, plastic pipes \| APOS, U | - GLO | market for polyvinylfluoride, film \| polyvinylfluoride, film \| APOS, U | - GLO |
| market for polymer foaming \| polymer foaming \| APOS, U | - GLO | market for water, ultrapure \| water, ultrapure \| APOS, U | - RoW |
| injection moulding \| injection moulding \| APOS, U | - CA- QC | market for dipropyl amine \| dipropyl amine \| APOS, U | - GLO |
| market for textile, jute \| textile, jute \| APOS, U | - GLO | market for textile, silk \| textile, silk \| APOS, U | - GLO |
| market for textile, kenaf \| textile, kenaf \| APOS, U | - GLO | market for synthetic rubber \| synthetic rubber \| APOS, U | - GLO |
| market for weaving, synthetic fibre \| weaving, synthetic fibre \| APOS, U | - GLO | market for textile, woven cotton \| textile, woven cotton \| APOS, U | - GLO |
| textile production, cotton, weaving \| textile, woven cotton \| APOS, U | - RoW | market for propylene glycol, liquid \| propylene glycol, liquid \| APOS, U | - RoW |
| market for glass tube, borosilicate \| glass tube, borosilicate \| APOS, U | - GLO | glass tube production, borosilicate \| glass tube, borosilicate \| APOS, U | - RoW |
| market for flat glass, coated \| flat glass, coated \| APOS, U | - RoW | market for chemical, organic \| chemical, organic \| APOS, U | - GLO |
| market for flat glass, uncoated \| flat glass, uncoated \| APOS, U | - RoW | market for tempering, flat glass \| tempering, flat glass \| APOS, U | - GLO |
| market for aluminium alloy, AlLi \| aluminium alloy, AlLi \| APOS, U | - GLO | market for folding boxboard carton \| folding boxboard carton \| APOS, U | - RoW |
| market for fibre, polyester \| fibre, polyester \| APOS, U | - GLO | market for steel, low-alloyed \| steel, low-alloyed \| APOS, U | - GLO |
| market for polypropylene, granulate \| polypropylene, granulate \| APOS, U | - GLO | market for transport, passenger car \| transport, passenger car \| APOS, U | - RoW |
| market for corrugated board box \| corrugated board box \| APOS, U | - CA- QC | market for waste plastic, mixture \| waste plastic, mixture \| APOS, U | - RoW |
| market for tap water \| tap water \| APOS, U | - RoW | market for waste glass \| waste glass \| APOS, U | - RoW |
| market for water, decarbonised \| water, decarbonised \| APOS, U | - US | market for wastewater, average \| wastewater, average \| APOS, U | - RoW |
| market for nylon 6 \| nylon 6 \| APOS, U | - RoW | market for nylon 6 \| nylon 6 \| APOS, U | - RoW |
| market for municipal solid waste \| municipal solid waste \| APOS, U | - RoW | market for waste polyurethane \| waste polyurethane \| APOS, U | - RoW |
| market for scrap aluminium \| scrap aluminium \| APOS, U | - RoW | market for blow moulding \| blow moulding \| APOS, U | - GLO |
| market for waste graphical paper \| waste graphical paper \| APOS, U | - RoW | market for waste packaging paper \| waste packaging paper \| APOS, U | - RoW |
| market for waste paperboard \| waste paperboard \| APOS, U | - RoW | market for waste polyethylene \| waste polyethylene \| APOS, U | - RoW |
| market for injection moulding \| injection moulding \| APOS, U | - GLO | market for carbon dioxide, liquid \| carbon dioxide, liquid \| APOS, U | - RoW |
| market for glass fibre \| glass fibre \| APOS, U | - GLO | market for paper sack \| paper sack \| APOS, U | - GLO |
| market for tissue paper \| tissue paper \| APOS, U | - GLO | market for kraft paper \| kraft paper \| APOS, U | - RoW |
| market for nylon 6-6 \| nylon 6-6 \| APOS, U | - RoW |  |  |

**Supplemental Table 3: Estimated impacts of individual colon cancer screening tests.** Means and 95% empirical confidence intervals estimated by Monte Carlo simulation (N=10,000)

|  | Greenhouse gas emissions (kg CO2 eq.) | | Damage to Human Health (DALY) | |
| --- | --- | --- | --- | --- |
| Screening Tests | Mean | Confidence Interval | Mean | Confidence Interval |
| FIT | 0.26894 | [0.25990, 0.27780] | 0.00219 | [0.00205, 0.00276] |
| Colo (incl. patient transport) | 39.60549 | [38.21661, 40.53643] | 0.32510 | [0.30992, 0.30992] |
| CTC (incl. patient transport) | 27.82841 | [26.44403, 28.73249] | 0.22303 | [0.21274, 0.24019] |
| Colo (incl. transport by all) | 43.29253 | [41.62843, 44.34857] | 0.35385 | [0.33715, 0.39957] |
| CTC (incl. transport by all) | 28.81548 | [27.35740, 29.75403] | 0.25178 | [0.23990, 0.27242] |

**Supplemental Table 4: Mean reductions in environmental impacts following transitions to electric vehicles.** Means and 95% empirical confidence intervals estimated by Monte Carlo simulation (N=10,000). Colonoscopy and CTC results correspond to the scenario where only patient transportation is modeled (provider and staff transportation are treated as fixed/unavoidable costs and thus not included in the model).

|  | Greenhouse gas emissions (kg CO2 eq.) | | Damage to Human Health (DALY) | |
| --- | --- | --- | --- | --- |
| Screening Strategies | Mean | Confidence Interval | Mean | Confidence Interval |
| FIT, 50% EVs | 31.41866 | [31.18133, 31.64572] | 0.26323 | [0.25586, 0.27580] |
| FIT, 100% EVs | 28.89949 | [28.63300, 29.14870] | 0.24809 | [0.23831, 0.26790] |
| Colonoscopy, 50% EVs | 38.60358 | [37.72658, 39.37454] | 0.32366 | [0.30763, 0.37524] |
| Colonoscopy, 100% EVs | 35.47017 | [34.45207, 36.29218] | 0.30479 | [0.28446, 0.40944] |
| CTC, 50% EVs | 57.77640 | [56.93831, 58.55178] | 0.47855 | [0.46395, 0.51716] |
| CTC, 100% EVs | 51.33307 | [50.38749, 52.17090] | 0.43997 | [0.41809, 0.51461] |

**Supplemental Figure 1: Estimated greenhouse gas emissions resulting from individual CRC screening tests**. 100,000 Monte Carlo simulations were performed for each test.

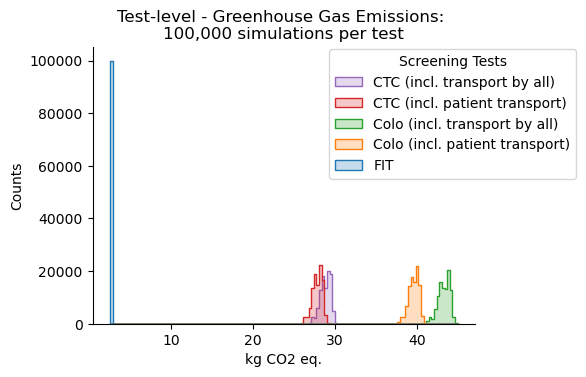

**Supplemental Figure 2: Estimated damage to human health resulting from performing individual CRC screening tests**. 100,000 Monte Carlo simulations were performed for each test.

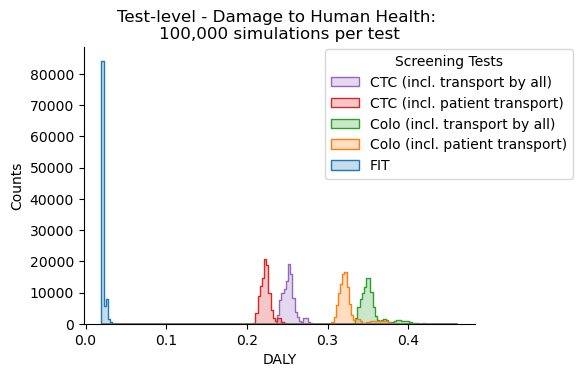

**Supplemental Figure 3: Projected greenhouse gas emissions resulting from different screening strategies over 10 years.** Orange and red bars correspond to scenarios where staff transportation is not considered an opportunity cost (e.g. assume that physicians, nurses, technicians and others would still travel from their homes to the medical center to perform other work, under a counterfactual scenario). The purple and green bars correspond to scenarios where patients and all staff do not travel to the medical center under a counterfactual scenario. Histograms summarize the results of 100,000 Monte Carlo simulations per screening strategy.

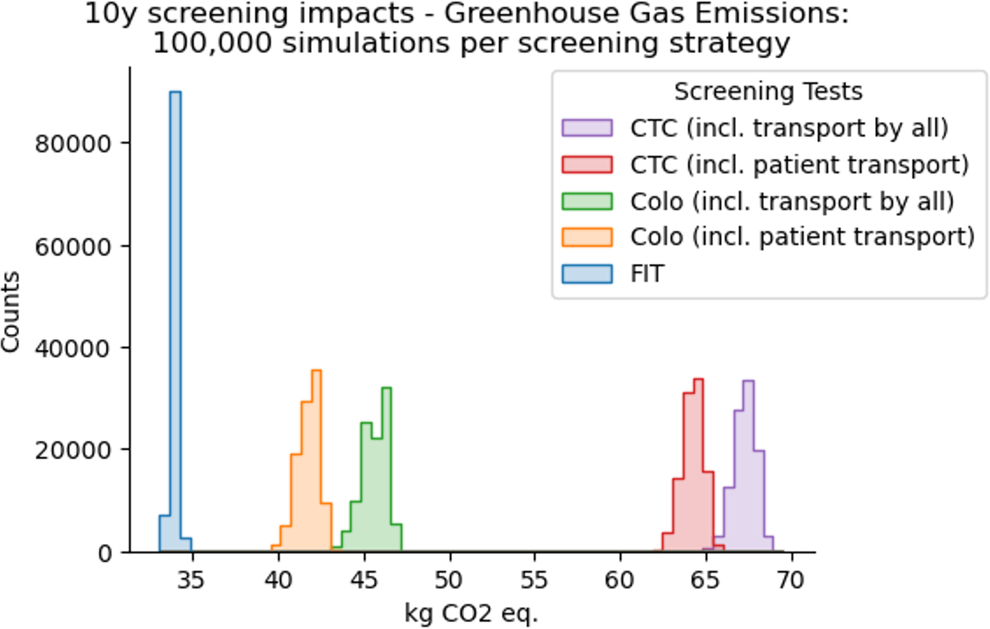

**Supplemental Figure 4: Projected damage to human health resulting from different screening strategies over 10 years.** Orange and red bars correspond to scenarios where staff transportation is not considered an opportunity cost (e.g. assume that physicians, nurses, technicians and others would still travel from their homes to the medical center to perform other work, under a counterfactual scenario). The purple and green bars correspond to scenarios where patients and all staff do not travel to the medical center under a counterfactual scenario. Histograms summarize the results of 100,000 Monte Carlo simulations per screening strategy.

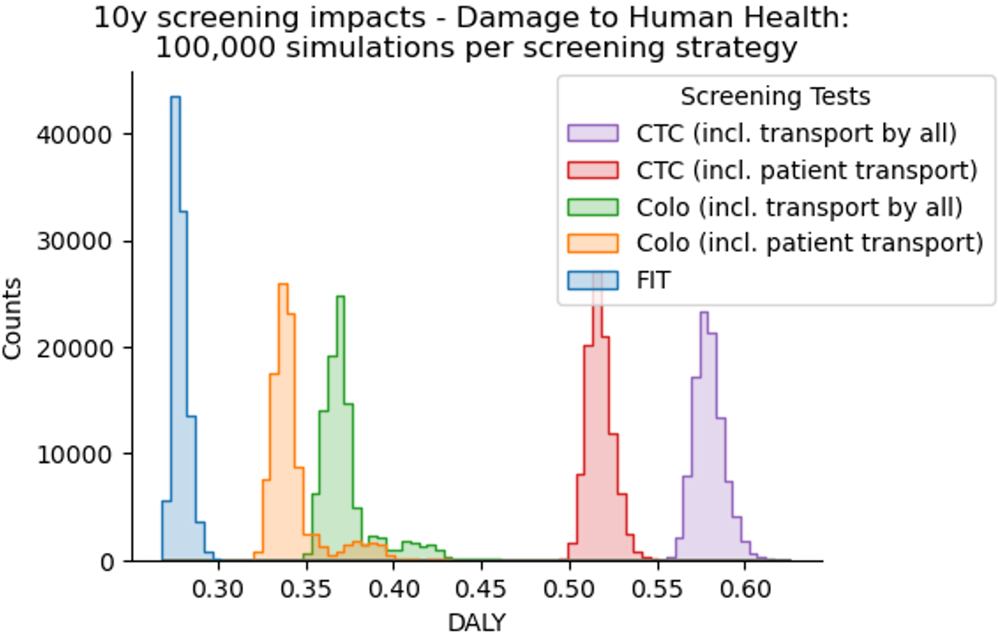
